# Precise perivascular space segmentation on magnetic resonance imaging from Human Connectome Project-Aging

**DOI:** 10.1101/2025.03.19.25324269

**Authors:** Yaqiong Chai, Hedong Zhang, Carlos Robles, Andrew Shinho Kim, Nada Janhanshad, Paul M Thompson, Ysbrand van der Werf, Eva M. van Heese, Jiyoung Kim, Eun Yeon Joo, Leon Aksman, Kyung-Wook Kang, Jung-Won Shin, Abigail Trang, Jongmok Ha, Emily Lee, Yeonsil Moon, Hosung Kim

## Abstract

Perivascular spaces (PVS) are cerebrospinal fluid-filled tunnels around brain blood vessels, crucial for the functions of the glymphatic system. Changes in PVS have been linked to vascular diseases and aging, necessitating accurate segmentation for further study. PVS segmentation poses challenges due to their small size, varying MRI appearances, and the scarcity of annotated data. We present a finely segmented PVS dataset from T2-weighted MRI scans, sourced from the Human Connectome Project Aging (HCP-Aging), encompassing 200 subjects aged 30 to 100. Our approach utilizes a combination of unsupervised and deep learning techniques with manual corrections to ensure high accuracy. This dataset aims to facilitate research on PVS dynamics across different ages and to explore their link to cognitive decline. It also supports the development of advanced image segmentation algorithms, contributing to improved medical imaging automation and the early detection of neurodegenerative diseases.

## Background & Summary

Perivascular spaces (PVS) are annular cerebrospinal fluid (CSF)-filled tunnels around blood vessels in the brain^1^. As conduits of the central nervous system (CNS) to the lymphatic drainage in the brain, PVS play a substantial role in the clearance function of the glymphatic (glial-lymphatic) system^2-4^.

Morphological changes in PVS have clinical implications, as enlarged PVS (ePVS) are linked to cerebrovascular diseases, including cerebral small vessel disease, cerebral amyloid angiopathy, hypertension, and stroke^3,5-8^. ePVS are associated with aging and cardiovascular risks in healthy adults as well as neurodegenerative diseases^9-11^. ePVS distribute throughout the white matter, basal ganglia, and hippocampus and are visualized using high-resolution Magnetic Resonance Imaging (MRI). Therefore, accurately identifying and segmenting ePVS on MRI is crucial for understanding their role in normal aging and brain diseases. ePVS have the same intensity characteristics as cerebrospinal fluid (CSF) on MRI, which is hypointense (dark) on T1-weighted (T1w) and hyperintense (bright) on T2-weighted (T2w) MRI images. Yet, they are varying in size from sub-millimeter to 3mm, and appear as thousands of thin, tubular structures. Further, ePVS are often confused with white matter hyperintensity and lacunar infarcts^12^, requiring high-level expertise that identifies ePVS in different shapes as round, ovoid, or tubular depending on the slice orientation of MRI^13^. These challenges make annotation difficult, resulting in limited datasets with ePVS labels.

Efforts have been made to explore various computational methods to segment ePVS, yet significant challenges remain. Earlier, Frangi filter-based method was adopted as it can detect ePVS without dependency on labeled data^14-16^. However, this results in low specificity, resulting in numerous false positives. More recently, supervised automated methods have demonstrated improved performance^17^ but often generate fraction of labeled ePVS, resulting in false negatives. This is partly due to the limited data available for training supervised models, preventing machine learning models from achieving accuracy for clinical utility. These limitations underscore the need for high-quality, comprehensive ePVS label datasets to improve the accuracy and reliability of segmentation methods for new subjects.

Here, we present a dataset of ePVS that has been finely segmented on T2w MRI. This dataset includes 200 subjects sampled from the Human Connectome Project Aging (HCP-Aging)^18^, ensuring a wide representation of ages and a uniform distribution across age groups ranging from 36 to 100 years old. We employ a combination of semi-supervised deep learning^19^, and iterative manual correction methods to label ePVS precisely. The release of this dataset aims to provide a reliable foundation for researchers to further study the dynamic changes of ePVS at a wide age range and investigate their potential link to various risk factors of cerebrovascular and neurodegenerative diseases, as well as cognitive decline. Additionally, it can be used to develop, train, and validate new image segmentation algorithms, thereby advancing the automation of medical imaging processing.

## Methods

### Subjects

This study leveraged data from the HCP-Aging Lifespan Release 2.0. At the time of recruitment, subjects were in good health, without a diagnosed history of neurologic or major psychiatric disorder, symptomatic stroke or Alzheimer’s disease. The dataset also includes the self-reported Pittsburgh Sleep Quality Index (PSQI) questionnaire^20^. Participants with Montreal Cognitive Assessment (MoCA) scores below 19 were excluded from the HCP-Aging^21^, but those with a MoCA with 20 or higher were included. However, it is noted that a MoCA score within the range of 18-25 is generally classified as mild cognitive impairment (MCI)^21^.

### Clinical characteristics

The patient selection workflow for the study dataset is illustrated in Fig. 1. Initially, there were 765 subjects with neuroimaging data available in HCP-Aging. First, we selected 217 subjects while considering as much as uniform distribution of age and sex. After visually confirming the image quality of T2w MRI, 17 were excluded due to image motion artifacts (n=11) or very large lacunar infarcts (n=6), resulting in 200 subjects in the following segmentation process (age: mean±SD=56.36±10.61 years, range=30-100 years).

**Figure 1.**
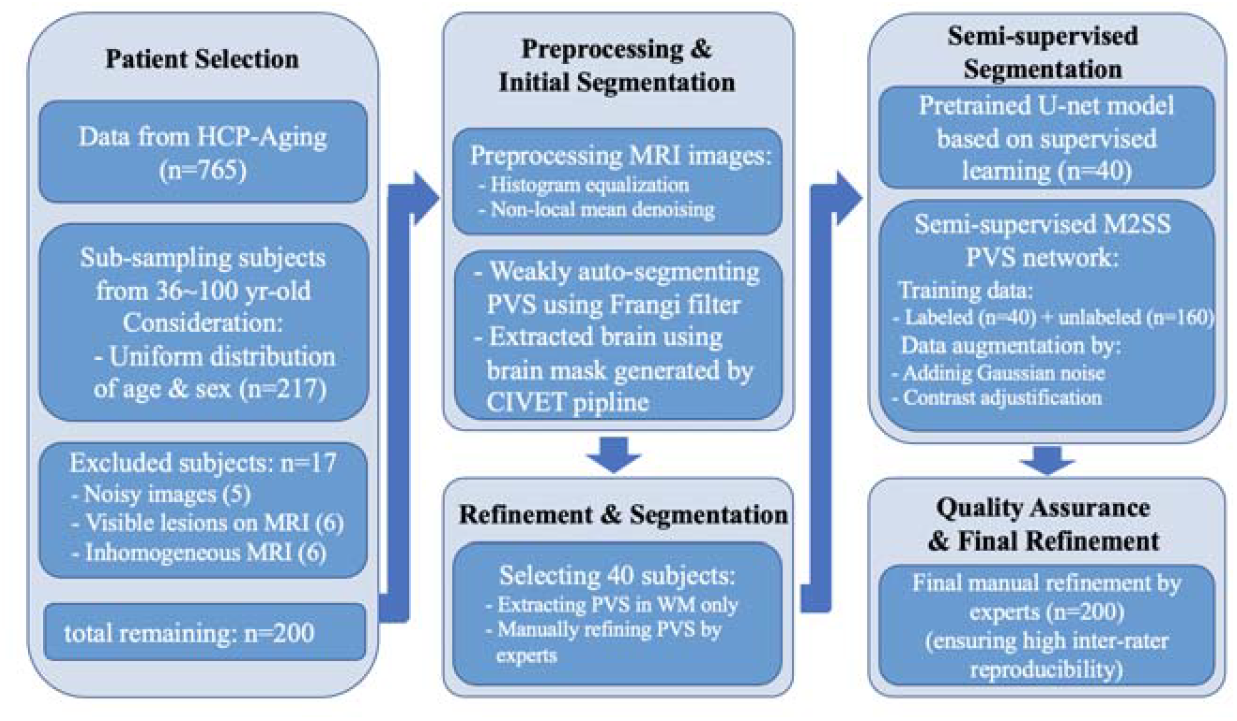
Work flow of generating high-quality PVS masks from Human Connectome Project Aging (HCP-Aging) dataset. M2SS: multi-scale semi-supervised PVS segmentation.

As the vascular risk factors are associated with ePVS, we present the distribution of study cohort’s body mass index (BMI), blood pressure, and comprehensive metabolic panel in Fig. 2. BMI was calculated from height (cm) and weight (kg) measurements collected during subject’s interviews using the formula weight (kg)/height (m)^2^. According to the Centers for Disease Control and Prevention (CDC)^22^, individuals with a BMI of 30 or higher are classified as obese. Systolic and diastolic blood pressure were measured with subjects in a seated position. According to the guidelines from the American College of Cardiology (ACC) and the American Heart Association (AHA)^23^, hypertension is defined as: a systolic blood pressure of 130 mmHg or higher, or a diastolic blood pressure of 80 mmHg or higher. High density lipoprotein (HDL), hemoglobin A1c (HbA1c), plasma glucose, triglycerides, vitamin D, and total cholesterol levels were tested from fasting blood samples. Diabetes is diagnosed using HbA1c and fasting glucose. According to the American Diabetes Association (ADA)^24^, diabetes is indicated by an A1c level of 6.5% or higher, or an fasting plasma glucose level of 126 mg/dL (7.0 mmol/L) or higher. The detailed measures and the diagnosis are shown in Table 1.

**Table 1.** Summary of the demographic, clinic, cognitive, and sleep quality measures of the patient population (n=200). Data are expressed as MEAN±SD unless otherwise specified.

|  | Value |
| --- | --- |
| Age [years (range)] | 61.3 (36-100) |
| Male sex [n (%)] | 87 (44) |
| Body mass index [kg/m<sup>2</sup>] | 26.6±5.2 |
| High-Density Lipoprotein | 60.9±17.2 |
| Hemoglobin A1c | 5.4±0.6 |
| Total cholesterol | 196.1±37.6 |
| Triglycerides | 108.1±68.7 |
| Glucose | 102.7±16.6 |
| 25-vitamin D | 34.2±15.3 |
| Systolic | 130.4±15.6 |
| Diastolic | 79.2±9.6 |
| Obesity [n (%)] | 42 (21) |
| Hypertension [n (%)] | 128 (64) |
| Diabetes [n (%)] | 9 (5) |
| Montreal Cognitive Assessment | 26.2±2.6 |
| Pittsburgh Sleep Quality Index | 4.5±2.4 |

**Figure 2.**
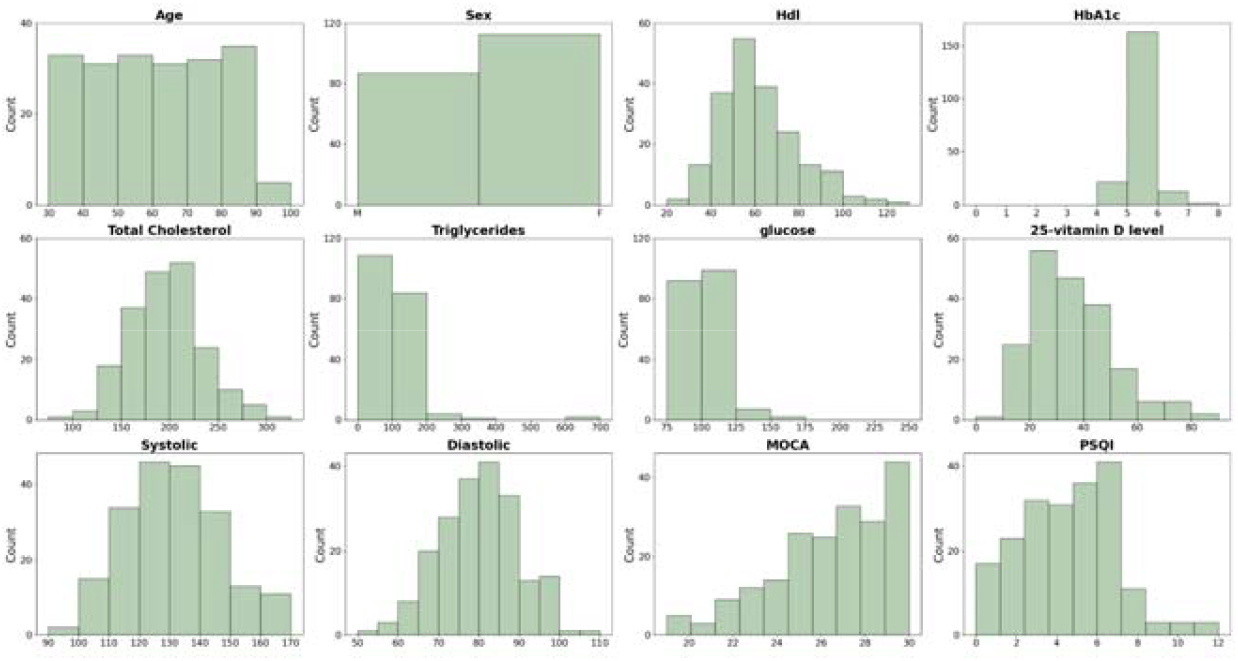
Demographic and clinical characteristics of the study subjects. HDL: high-density lipoprotein; HbA1c: hemoglobin A1c; MoCA: Montreal Cognitive Assessment; PSQI: Pittsburg sleep quality index.

### Imaging parameters

Subjects were scanned using Siemens 3T Prisma scanner with an 80 mT/m gradient coil and a 32-channel Siemens receive head coil, at Washington University in St. Louis, Missouri. T1 multi-echo MP-RAGE scans were acquired with the following acquisition parameters: repitition time (TR) = 2500ms, time of inversion (TI) = 1000ms, time to echo (TE) = 1.8/3.6/5.4/7.2 ms, 4 echoes per line of k-space, voxel size = 0.8 mm isotropic, FOV=256 × 240 × 166 mm, matrix = 320 × 300 × 208 slices, flip angle =8°. T2 turbo-spin echo (TSE) scans were acquired with TR/TE = 3200/564 ms, turbo factor = 314, and same voxel size, FOV and matrix with T1 images.

### Image processing

T1 images were preprocessed using Montreal Neurological Institute (MNI)-CIVET pipeline^25^, including brain-extraction and bias correction. Both T1 and T2 images were linearly co-registered together, to eliminate the slight slice misalignment during image acquisition. Binary brain mask, excluding the dura matter, cerebellum, and brainstem, produced by CIVET pipeline were applied to T2w images^25^. Then histogram equalization was performed on T2w images, making ePVS more discernible^26^. Non-local mean denoising was employed to remove noise from T2w images^27^.

### Initial weakly auto-segmentation of ePVS

We performed auto-segmentation using the Frangi filter. The parameters for the Frangi filter were empirically selected to accentuate tubular shapes across a range of diameters, using sigma values of [0.01, 0.05, 0.1, 0.25, 0.5, 0.75, 1.0]. Given that ePVS typically span only 1-2 voxels in diameter, these sigma values were chosen to be smaller than those typically used for vascular segmentation, aiming to detect finer tubular shapes. The smaller sigma values are more noise-sensitive, highlighting the necessity of prior denoising. The MNI-CIVET pipeline was utilized to apply a white matter (WM) mask, effectively isolating ePVS within the WM by removing false positives outside WM^25^.

### Manual refinement for 40 subjects

After applying the Frangi filter, the initial segmentation underwent manual refinement, which was performed using ITK-snap^28^. To ensure the training samples for the upcoming semi-supervised learning, 40 subjects were selected out of the initial 200 subjects, by selecting four to five subjects from each decade of life. Two neuroanatomists manually refined them, aiming to correct and complete the PVS segmentation flagged by the Frangi filter and eliminate mis-labeled PVS.

Given the widespread distribution of ePVS throughout the brain, the white matter including the centrum semiovale (CS) and the deep gray matter including the basal ganglia (BG) and hippocampus are known to exhibit the highest PVS burden^7,29^. In this study, we focus specifically on ePVS located within the white matter. Consequently, our neuroanatomists did not include ePVS in the deep gray matter region.

When correcting ePVS and adding new ePVS for completeness, the primary criterion used was the appearance across the 3 orthogonal slices (i.e. axial, coronal, and sagittal). In one slice, most ePVS appeared as long tubular structures, whereas on the other views, they appeared as small ovoid shapes, ranging from submillimeter (one voxel thick on HCP-A T2w images) to 3 mm in diameter^30^. To evaluate the tubular shape of ePVS more specifically on T2w MRI with 0.8mm isotropic resolution, we examined whether hyperintense voxels consisting of an ovoid shape were present on at least three consecutive slices in one view and linearity in voxels appearing on 1-2 slices in other views. However, some ePVS that were located obliquely to all the 3 orthogonal planes appeared with a shorter tubular shape in all 3 views^13^. The neuroanatomists confirmed these structures by simultaneously examining the 3 orthogonal slices to differentiate them from lacunes, exhibiting a hyperintense “donut” shape, and white matter hyperintensities (WMH), exhibiting oval shape in all the 3 orthogonal views. These features are often confused with or connected to ePVS when evaluated solely on a single view^31^.

Automated segmentation generated by Frangi filter often misidentified WMH and lacunes as ePVS, particularly when these lesions are adjacent or connected to ePVS^29,31^. To address this issue, neuroanatomists manually excised voxels corresponding to WMH and lacunes by applying an imaginary boundary considering the tubular shape of ePVS.

### Semi-supervised deep learning segmentation

The manually refined labels from the 40 selected subjects above were used to train and validate a U-Net model for supervised learning. The U-Net model was designed with a five-layer encoder-decoder structure; each encoder layer consists of two sets of 3×3 convolutional layers and Leaky ReLU layers, followed by a MaxPooling layer^19^. The loss function combined Dice loss and binary cross-entropy loss to address the issue of class imbalance present in the dataset, where ePVS occupy a significantly smaller proportion of the images. This choice of loss function improved model accuracy in imbalanced samples. Unlabeled images (n=40 randomly chosen from 160 data) were augmented by enhancing contrast and adding noise^32^, aiming to enable the model to capture various curved and subtle ePVS with intensity levels closer to the surrounding white matter. These augmented data were included in the final step for semi-supervised learning process. Dice discrepancies between the model’s predictions for augmented images and their original counterparts were used to calculate consistency loss, thus informing model updates. This strategy ensured that the model was exposed to both labeled and unlabeled dataset, accommodating the variability in ePVS appearances. After training, the model predicted ePVS segmentation across the left 160 dataset (aside 40 from initial 200).

### Quality assurance and final refinement

Manual refinement of the deep learning-generated ePVS labels was performed by the aforementioned two neuroradiologists and one medical student trained by them, following the same protocol described earlier. To ensure consistency and accuracy, the three members convened weekly to reach a consensus on any uncertain cases. Fig. 3-5 present the final refined ePVS annotations on T2w MRI, along with 3D renderings of the PVS burden for three subjects representing ages in 30’s, 50’s and 70’s.

**Figure 3.**
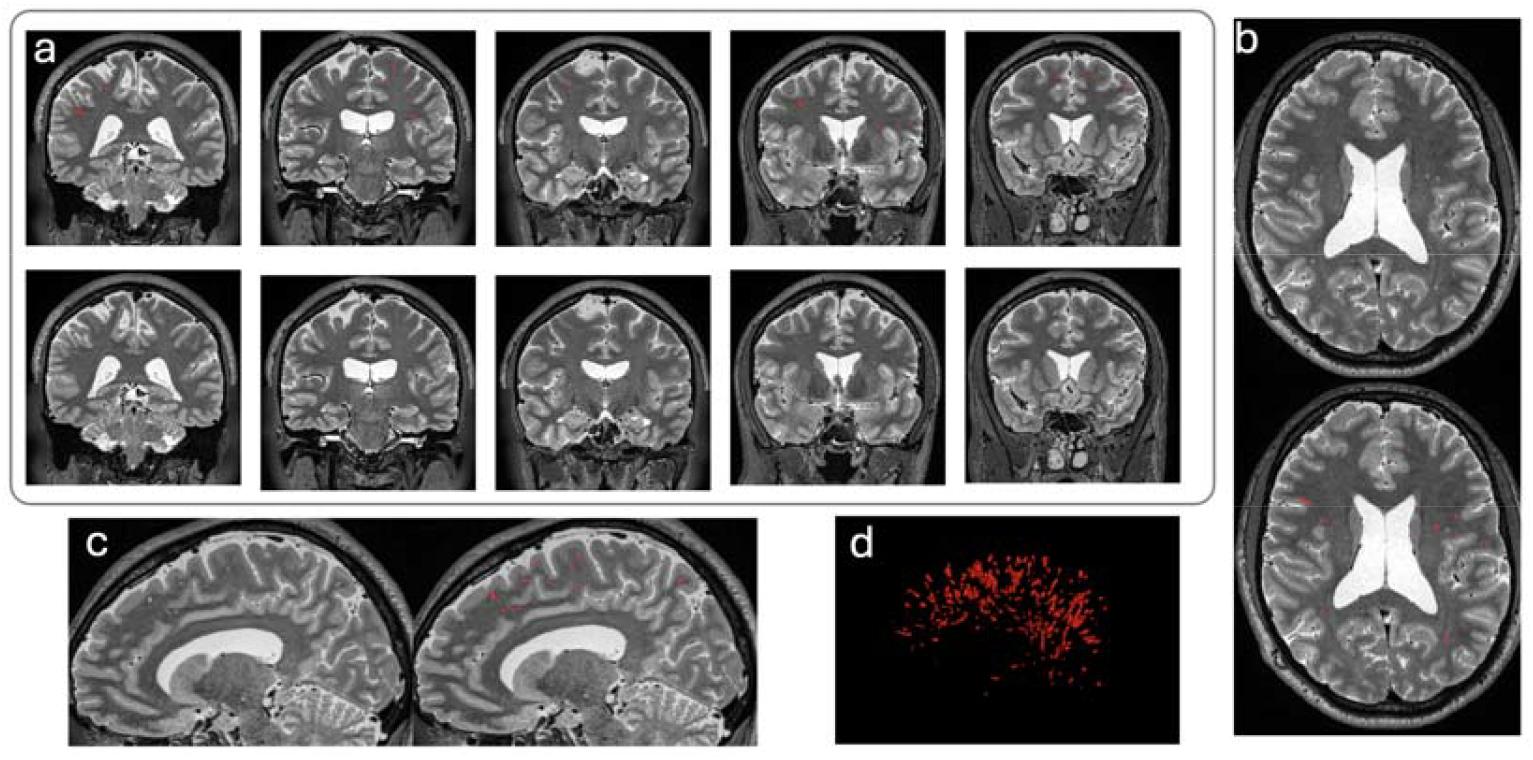
T2 MRI with PVS (red) of a 36-yr old male subject, in coronal view (a), axial view (b), and sagittal view (c). Panel (d) shows the rendered 3D PVS map.

**Figure 4.**
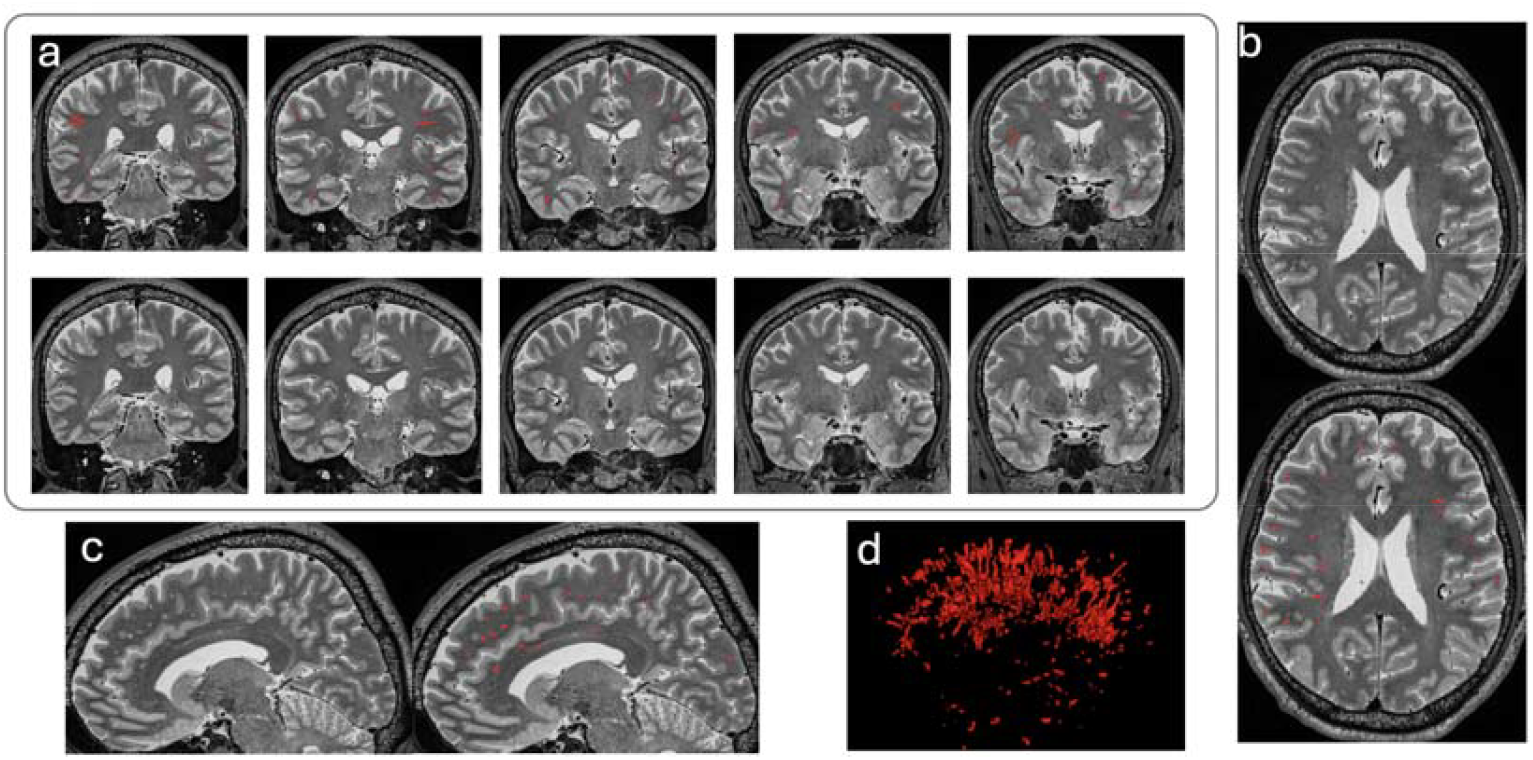
T2 MRI with PVS (red) of a 56-yr old male subject, in coronal view (a), axial view (b), and sagittal view (c). Panel (d) shows the rendered 3D PVS map.

**Figure 5.**
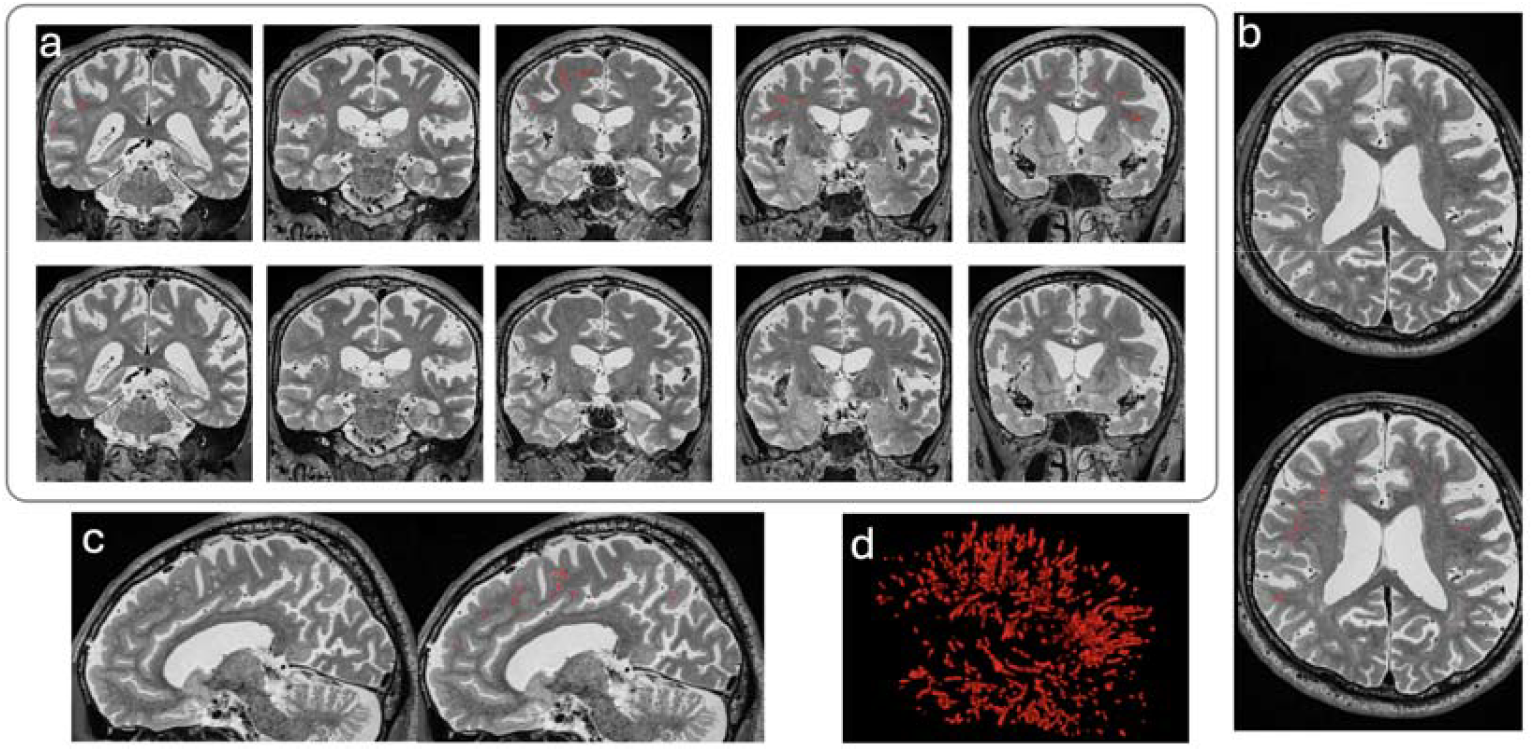
T2 MRI with PVS (red) of a 76-yr old male subject, in coronal view (a), axial view (b), and sagittal view (c). Panel (d) shows the rendered 3D PVS map.

## Data Availability

Following NIMH Data Archive (NDA) derived data sharing policies, the raw T2w MRI images of the 200 subjects ePVS labels were shared in BALSA database at https://balsa.wustl.edu/study/x9Z66 as well as https://openneuro.org/datasets/ds005595/versions/1.0.0
CIVET was used to pre-process T1w images. https://github.com/aces/CIVET_Full_Project.

## Data Records

All HCP-A imaging data could be downloaded at NIMH DATA ARCHIVE (NDA). Data were provided [in part] by the Human Connectome Project, WU-Minn Consortium (Principal Investigators: David Van Essen and Kamil Ugurbil; 1U54MH091657) funded by the 16 NIH Institutes and Centers that support the NIH Blueprint for Neuroscience Research; and by the McDonnell Center for Systems Neuroscience at Washington University. Metabolic panels, vitals including BMI and blood pressure, PSQI, and MoCA are available as text files. The clinical characteristics are available as a single Microsoft Excel spreadsheet. T2w images are provided in de-identified NIFTI format, and PVS segmentations are binarized masks for each subject.

## Technical Validation

To validate the consistency and accuracy of the segmentation results, we randomly chose 10 cases, and evaluated the reproducibility across three raters. We used Fleiss Kappa statistics to calculate the inter-rater reliability among the three raters. As an additional agreement metric, we computed the DICE agreement coefficient between all pairs of experts. The Kappa statistics and DICE similarities are presented in Table 2.

**Table 2.**
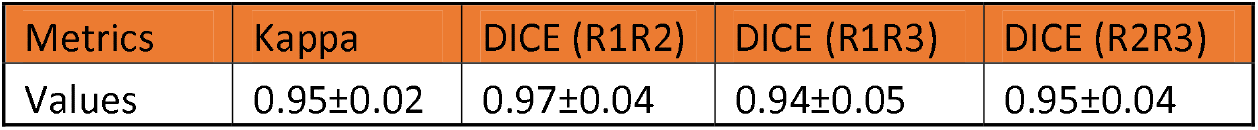
Summary of Kappa and DICE similarity index among three raters. R1, R2 and R3 indicates three independent raters.

## Code Availability

Following NIMH Data Archive (NDA)’s derived data sharing policies, the 200 subjects’ raw T2w MRI images ePVS labels were shared in BALSA database: https://balsa.wustl.edu/study/x9Z66 as well as https://openneuro.org/datasets/ds005595/versions/1.0.0

CIVET was used to pre-process T1w images (https://github.com/aces/CIVET_Full_Project). The pre-segmentation code using Frangi filter and semi-supervised learning codes can be found on our Github website (https://github.com/Yaqiongchai/PVS_segmentation). The refinement were generated using ITK-SNAP version 3.8.0.

## Acknowledgements

This work was supported by the following grants: National Institute on Aging (P30-AG066530-03), the Foundation of the ASNR Grants (AWD-00008862), NIA/California Institute for Research and Education-(NCIRE)/UCSF, WEI2990-18 (#U01 AG024904), and Micheal J Fox Foundation -The Parkinson’s Progression Markers Initiative (#MJFF-023385). Dataset used in this publication was supported by the National Institute on Aging of the National Institutes of Health under Award Number U01AG052564. Research reported in this publication was supported by the Office Of The Director, National Institutes Of Health of the National Institutes of Health under Award Number S10OD032285. The content is solely the responsibility of the authors and does not necessarily represent the official views of the National Institutes of Health.

## Author contributions

Study design, Y. C. and H.K.; statistical analysis, H. Z., C.R, and A.S.K.; manual refinement, A.S.K., K.W.K, and J.K.; and manuscript drafting or manuscript revision for important intellectual content, all authors.

